# Social clustering of unvaccinated children: measles-mumps-rubella vaccination coverage in schools in the Netherlands

**DOI:** 10.1101/2022.05.12.22273875

**Authors:** Don Klinkenberg, Albert Jan van Hoek, Irene Veldhuijzen, Susan Hahné, Jacco Wallinga

## Abstract

**Background:** For the measles-mumps-rubella (MMR) vaccine, the WHO-recommended coverage for herd protection is 95% for measles and 80% for rubella and mumps. However, a national vaccine coverage does not reflect social clustering of unvaccinated children, e.g. in schools of Orthodox Protestant or Anthroposophic identity in The Netherlands. To fully characterise this clustering, we estimated one-dose MMR vaccination coverages at all schools in the Netherlands.

**Methods:** By combining postcode catchment areas of schools and school feeder data, each child in the Netherlands was characterised by residential postcode, primary and secondary school (referred to as school career). Postcode-level vaccination data were used to estimate vaccination coverages per school career. These were translated to coverages per school, stratified by school identity.

**Results:** Most schools had vaccine coverages over 99%, but major exceptions were Orthodox Protestant schools (63% in primary and 58% in secondary schools) and Anthroposophic schools (67% and 78%).

**Conclusions:** School-level vaccine coverage estimates reveal strong clustering of unvaccinated children. The school feeder data reveal strongly connected Orthodox Protestant and Anthroposophic communities, but separated from one another. This suggests that even at a national one-dose MMR coverage of 97.5%, thousands of children per cohort are not protected by herd immunity.

## Introduction

The impact of childhood vaccination programmes on protecting children from infection can be measured in various ways. Common indicators are the remaining disease incidence and disease burden, and the national vaccination coverage [1]. For measles, WHO recommends a threshold value for the vaccination coverage of 95% for two doses [2], and for rubella and mumps of 80% [3]. Many EU countries have an explicit target coverage for the measles-mumps-rubella (MMR) vaccine of 95% [1]. However, outbreaks occur among unvaccinated children when they are clustered, even at high values for an average national vaccination coverage [4]. The Netherlands, in particular, have an average national vaccination coverage for MMR that is among the highest in Europe (http://data.euro.who.int/cisid/), but the substantial socio-geographic clustering of unvaccinated children results in repeated outbreaks of measles, mumps and rubella [5-7].

Clustering of unvaccinated children is often described geographically [8-13], but most contacts between children are made at school [14]. Indeed, daycare centres or schools play a major role during outbreaks in well-immunised countries [15-18]. In the Netherlands, the clustering of unvaccinated children at school is enhanced by a school system where parents are free to start and choose schools based on their own philosophy or conviction, resulting in a range of registered school identities. There are two social groups with their own school identities that are known for a lower vaccination coverage: Orthodox Protestants, who live spatially clustered in the so-called Bible Belt and have a vaccination coverage of about 60% [19, 20]; and Anthroposophics, who live spatially more dispersed and have school vaccination coverages ranging from 60% to 90% [21]. There is also a more diffuse group of people who fear side-effects and perceive the diseases as mild [22], but there is not one school identity linked to that group.

Vaccination coverages by school are routinely collected in various countries, including Germany and the United States [17, 18, 23-25], but reporting is frequently done by averaging school coverages locally, by county or district [24, 26]. Whereas this is very helpful for signaling which regions are at risk of infection, an average or even a local average does not fully capture the variability between schools, which is essential to fully understand the risk of outbreaks [25]. For that, we require the complete frequency distribution of school vaccination coverages.

The aim of our study was to estimate the MMR vaccination coverages for all primary and secondary schools in the Netherlands, and assess the variation in vaccination coverage in primary and secondary schools. We focussed on a single-dose rather than the WHO-recommended two-dose vaccination, because the second dose in the Netherlands is given only at nine years of age so that the one-dose coverage is a better indication of protection in primary schools. Also, a single-dose coverage better reflects willingness versus refusal to vaccinate, and shows where the completely unvaccinated children are. We discuss the implications of the observed variation for the risk of outbreaks in a population with high mean vaccination coverage.

## Methods

Briefly, we assumed a steady state population of children characterised by residential postcode, primary school, and secondary school (school career). This population is constructed by use of school-level catchment data of the postcode areas in which school children live, and school feeder data of children moving from primary to secondary school. The steady-state assumption allowed us to use postcode-level coverage data for a single cohort to estimate coverages in both primary and secondary schools. We used a statistical hierarchical model, in which the vaccination status of a child is explained by the school career, with a fixed effect of the school identities (identity career) and random effects of the two schools. The school feeder data create dependencies of coverage estimates between primary and secondary schools sharing many children, which is especially relevant if those schools have different identities.

### Datasets

We used five datasets to estimate school vaccination coverages (dark green in Figure 1), and two datasets for validation of estimated coverages (Utrecht and Anthroposophic school data).

**Figure 1.**
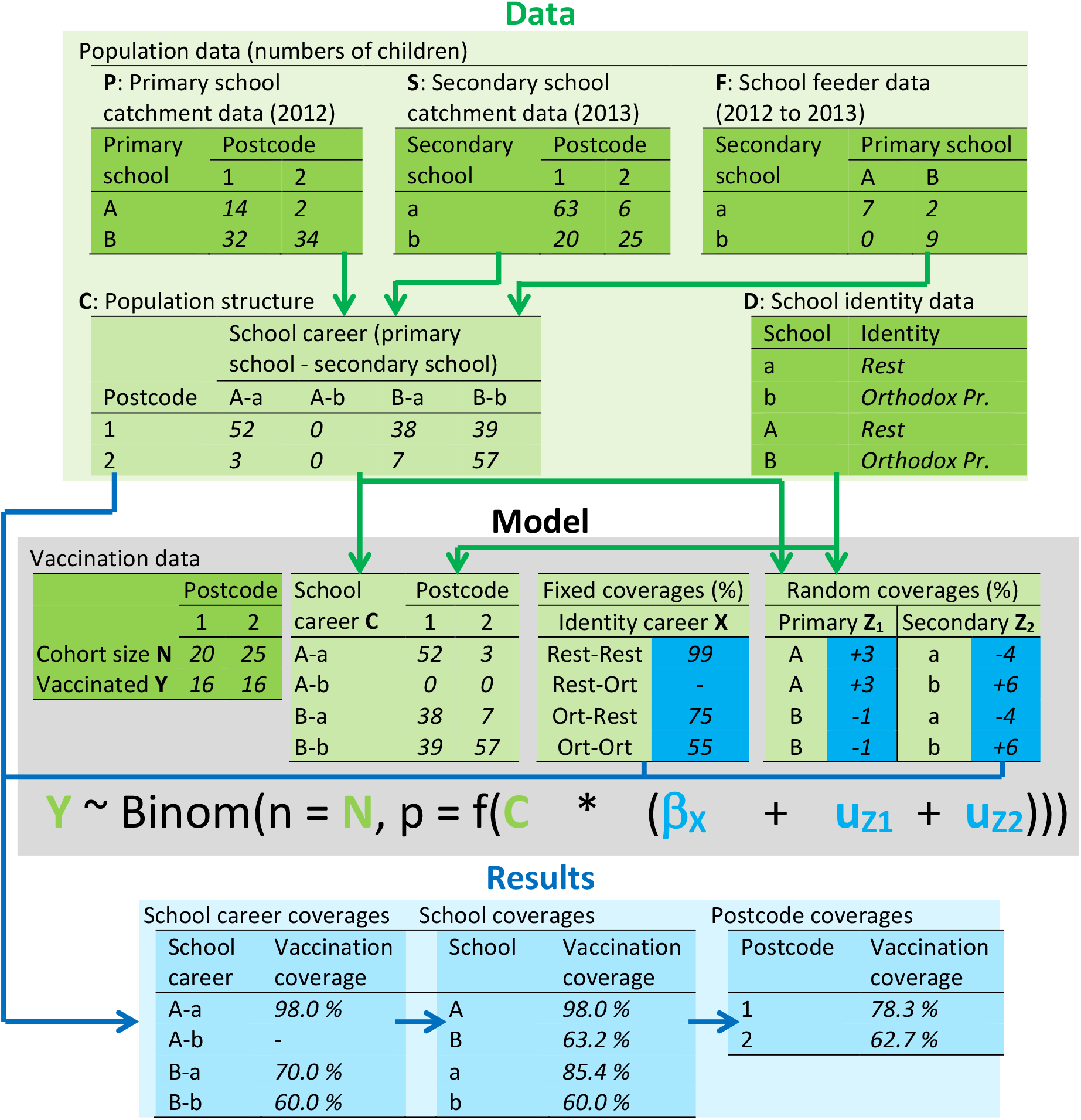
Conceptual outline of the method to estimate school vaccination coverages (full details in Supplement S1). Numbers are fictional but mutually consistent. Green tables are data: dark green are original datasets, light green are linked data. Blue tables are results: dark blue are original estimates (logit-transformed in the actual analysis), light blue are derived quantities. Top (data): three population datasets are linked to build the population structure of numbers of children by school career and residential postcode area. Middle (model): regression model with vaccination data per postcode as dependent variable; the independent variables are the school careers of the children, mapped to postcode areas through function f(.), in which the population structure is used to map school career coverages to postcode area coverages, and in which the logit(coverage) of each school career is modelled as the sum of a fixed identity career effect and two random school effects. Bottom (results): the estimated coverages and deviations are used to calculate the coverage per school career, and combined with the population structure to calculate the coverages per school and per postcode.

For estimation, we combined postcode-level MMR vaccination coverage data with postcode-level data of the schools that children attend (primary school and secondary school catchment data), school-level data on numbers of children moving from primary to secondary schools (school feeder data), and school identity data. The coverage data came from the Dutch vaccination registry Praeventis [27], of which we used the single MMR vaccination status in 2013 of 197,382 children of birth cohort 2003 at age 10. In the Netherlands, MMR is given at 14 months and 9 years of age, so focusing on a single-dose vaccination better reflects vaccine coverage across all ages in schools. The school catchment, feeder, and identity data came from the Education Executive Agency of the Ministry of Education [28], of which we used primary school catchment data from October 2012, secondary school catchment data from October 2013, and feeder data from September 2013.

Schools were categorised into 15 groups based on their identity [28]. Most identities are related to religion, but exceptions are Municipal (“Openbaar”) which are run by the local government, and General (“Algemeen bijzonder”) which is a heterogeneous group of schools with various non-religious backgrounds (e.g. Montessori, Dalton, Jenaplan). Special types of identity were “Collaborations”, which consisted of schools categorised as collaboration (“Samenwerking”) of two or more identities among Protestant, Roman Catholic, General, and/or Municipal; and “Other”, which consisted of all schools categorized as Rest (“Overige”) in the dataset, or left blank.

For validation we used data collected by the municipal health service Utrecht (Utrecht data), which covers part of the Bible Belt and consists of actual coverage data for 488 schools; and data from a study in 2014 on vaccination hesitancy on Anthroposophic schools (Anthroposophic school data), consisting of coverage data for 11 schools [21].

### Statistical analysis

Figure 1 shows how the data are used in a model to obtain the MMR coverage per school (more detail in Supplement S1). First, the school catchment data and feeder data were combined to build the population structure, in which each child is characterized by a postcode and a school career, i.e. a primary and secondary school, one of which the child actually attends and the other which it has attended or will attend. Then, this population structure is used with the vaccination data and school identities in a Bayesian regression analysis in stan (mc-stan.org), called from R statistical software [29] with the Rstan package (mc-stan.org/rstan). Conceptually, through the regression equation (Figure 1) the observed coverages in postcode areas are explained by the school careers of the children living in that postcode area, with a fixed effect due to the identities of the schools, and a random effect for each school.

The regression analysis resulted in estimated vaccination coverages per school career, which were used to derive our results of interest: vaccination coverages per school. The analysis also provided estimated coverages per postcode area.

Because there were 15 different school identities in the data, and therefore many more possible identity careers, we had to aggregate identities and identity careers to limit the number of variables in the regression. This was done in two preliminary analyses. The first was to define a “Rest” identity by grouping all identities without evidence of lower coverage than the major identities (Municipal, Protestant, and Roman Catholic). That resulted in five primary school identities and three secondary school identities. The second was to aggregate identity careers where possible, resulting in a final set of eight identity careers in the final analysis (see Supplement S1 for more detail).

## Results

### Aggregating school identities and identity careers

We first ran two preliminary regression analyses (detail in Supplement S1) in order to reduce the number of school identities and identity careers in our final analysis. Most school identities were aggregated into one “Rest” group, but identities that remained separate were Anthroposophic and Orthodox Protestant in both primary and secondary schools, and General and Hindu in primary schools (Table 1). From these remaining identities, 15 identity careers were formed, of which eight were kept separate after a second aggregation step to obtain our final results (Table 2).

**Table 1.** Identities of primary and secondary schools: numbers of schools per original identity, and identity after aggregation

| Identity | Primary schools |  | Secondary schools |  |
| --- | --- | --- | --- | --- |
|  | *n* | Aggregated identity | *n* | Aggregated identity |
| Municipal | 2444 | Rest | 352 | Rest |
| General | 497 | General | 392 | Rest |
| Anthroposophic | 70 | Anthroposophic | 11 | Anthroposophic |
| Roman Catholic | 2236 | Rest | 248 | Rest |
| Protestant | 1882 | Rest | 281 | Rest |
| Reformed (Liberated) | 118 | Rest | 9 | Rest |
| Orthodox Protestant | 180 | Orthodox Protestant | 34 | Orthodox Protestant |
| Evangelic | 13 | Rest | 3 | Rest |
| Evangelical Fraternity | 2 | Rest | 0 | - |
| Hindu | 6 | Hindu | 0 | - |
| Islamic | 43 | Rest | 1 | Rest |
| Jewish | 2 | Rest | 2 | Rest |
| Interconfessional | 12 | Rest | 4 | Rest |
| Collaborations | 65 | Rest | 362 | Rest |
| Other | 0 | - | 71 | Rest |
| Total | 7570 |  | 1770 |  |

**Table 2.** Identity careers: numbers of children in feeder data, and aggregate careers after the second aggregation step (Ant = Anthroposophic, Gen = General, Hin = Hindu, Ort = Orthodox Protestant, Rest = Rest)

| Primary school |  |  | Secondary school identity |  |  |  |
| --- | --- | --- | --- | --- | --- | --- |
| identity | Anthroposophic |  | Orthodox Protestant |  | Rest |  |
|  | *n* | Aggregate career | *n* | Aggregate career | *n* | Aggregate career |
| Anthroposophic | 679 | Ant - Ant | 0 | - | 876 | Ant - Rest |
| Orthodox | 0 | - | 4298 | Ort - Ort | 765 | Ort - Rest |
| Protestant |  |  |  |  |  |  |
| Hindu | 0 | - | 0 | - | 262 | Hin - Rest |
| General | 182 | Gen - Rest | 3 | Rest - Ort | 13246 | Gen - Rest |
| Rest | 778 | Rest - Rest | 782 | Rest - Ort | 182471 | Rest - Rest |

### Estimating school vaccination coverages

The overall vaccination coverage in our dataset (one dose of MMR vaccine in birth cohort 2003 at the age of 10) was 97.5%. Figure 2 shows the comparison of estimated school vaccination coverages with the validation datasets. In all Utrecht schools as well as the Anthroposophic schools there is a clear correlation between true and estimated coverage, and almost all credible intervals include the true coverage. Two Rest schools with coverages below 90% have too high estimates; these are both Protestant schools.

**Figure 2:**
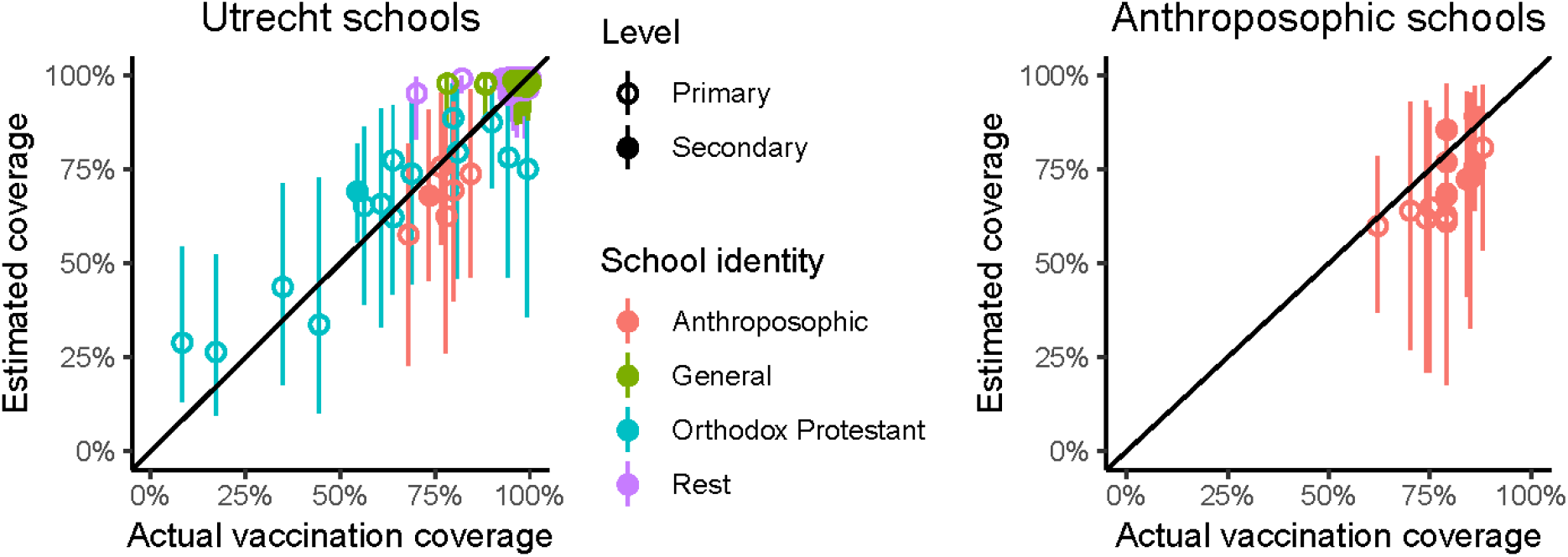
validation plots of estimated vaccination coverages with 95% credible interval compared to actual vaccine coverage in Utrecht, 2013 (left) and on Anthroposophical schools in the Netherlands, 2012 (right).

There is clear spatial clustering of unvaccinated children: most postcode areas had vaccination coverages around 99%, but about 4% of postcode areas had coverages below 95%, mainly in the Bible belt (Figures 3, 4). At school level, nonetheless, clustering was much more pronounced. The large majority of schools have have higher coverages than postcode area, above 99%, whereas specific schools have much lower coverages. In primary schools, vaccination coverage on Orthodox Protestant schools (62.8%) is slightly lower than on Anthroposophic schools (66.6%), whereas in secondary schools this difference is much larger (58.4% and 78.4%, respectively, Table 3). General primary schools have a slightly lower vaccination coverage than Rest primary schools (Table 3), but less than 1% of all these schools have a coverage below 95% (Figure 3). Coverages of Hindu schools are estimated to be lower than in the Rest or General schools, but this result is highly uncertain because of the small number of schools (Table 3). Of all identities, Orthodox Protestant schools have the largest variation in vaccine coverage (Figure 3).

**Table 3.** Vaccination status by school identity: mean vaccination coverage at school level (95% credible interval), and number of unvaccinated children per cohort of 197,382 (95% credible interval across all schools).

| School identity | Primary schools |  | Secondary schools |  |
| --- | --- | --- | --- | --- |
|  | Mean vaccination | Unvaccinated | Mean vaccination | Unvaccinated |
|  | coverage per school | children per cohort | coverage per school | children per cohort |
| Anthroposophic | 66.6% [59.6 ; 73.4] | 496 [402 ; 588] | 78.4% [71.3 ; 85.5] | 242 [164 ; 321] |
| Orthodox | 62.8% [60.3 ; 65.2] | 1904 [1799 ; 2009] | 58.4% [52.5 ; 64.0] | 1958 [1853 ; 2063] |
| Protestant |  |  |  |  |
| Hindu | 74.5% [50.3 ; 94.6] | 60 [15 ; 95] |  |  |
| General | 97.4% [96.4 ; 98.1] | 340 [243 ; 448] |  |  |
| Rest | 98.7% [98.6 ; 98.8] | 2198 [2041 ; 2366] | 98.4% [98.3 ; 98.5] | 2797 [2655 ; 2942] |
| All schools | 97.5% [97.4 ; 97.5] | 4998 [4872 ; 5129] | 97.5% [97.3 ; 97.7] | 4998 [4872 ; 5129] |

**Figure 3:**
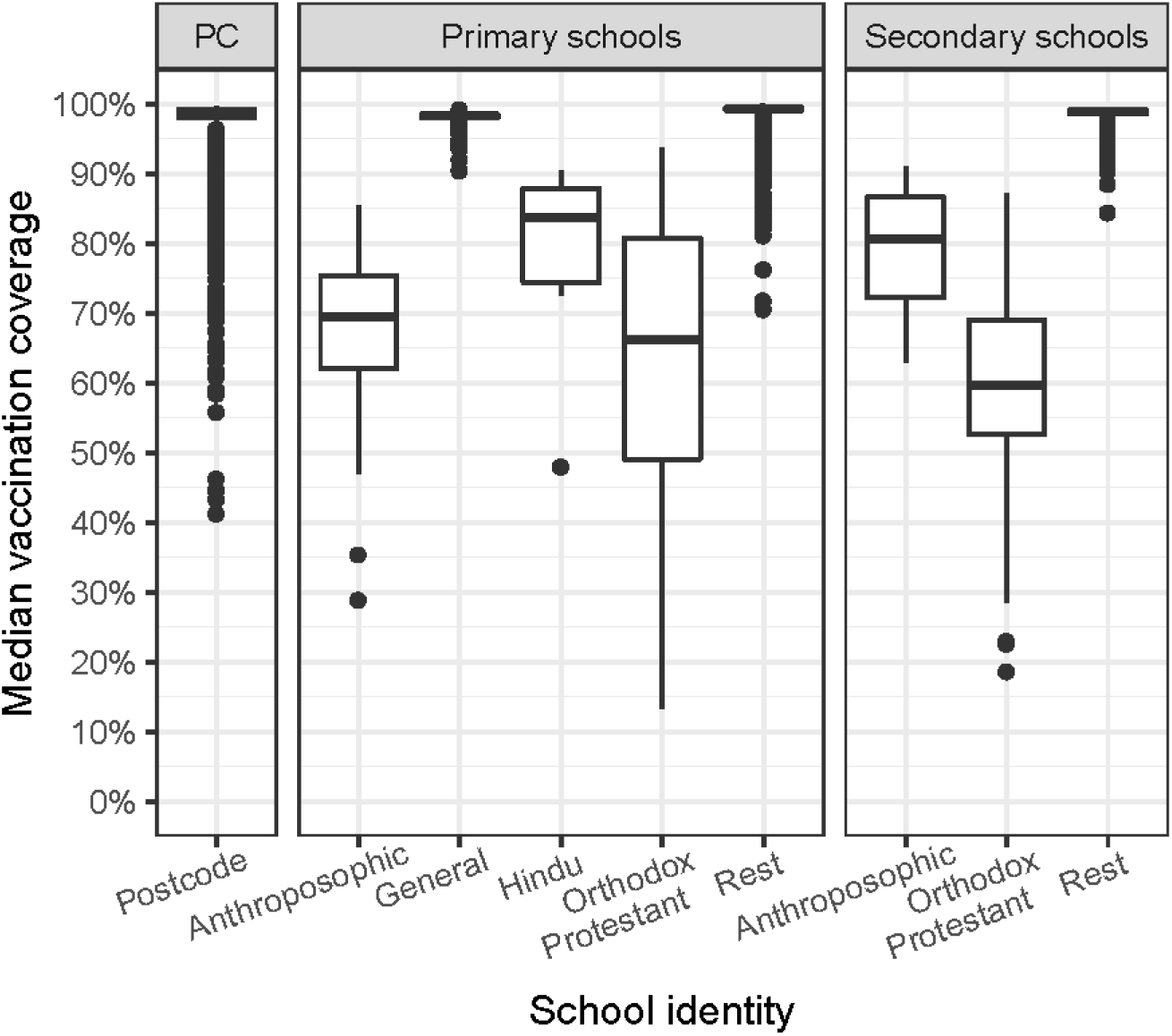
posterior median vaccination coverages of all schools, grouped by school identity. Displayed are medians (of the median coverages) with interquartile range of (IQR, boxes), and whiskers extending to 1.5 times the IQR or most extreme value, whichever is closest to the median. Dots are outliers.

**Figure 4:**
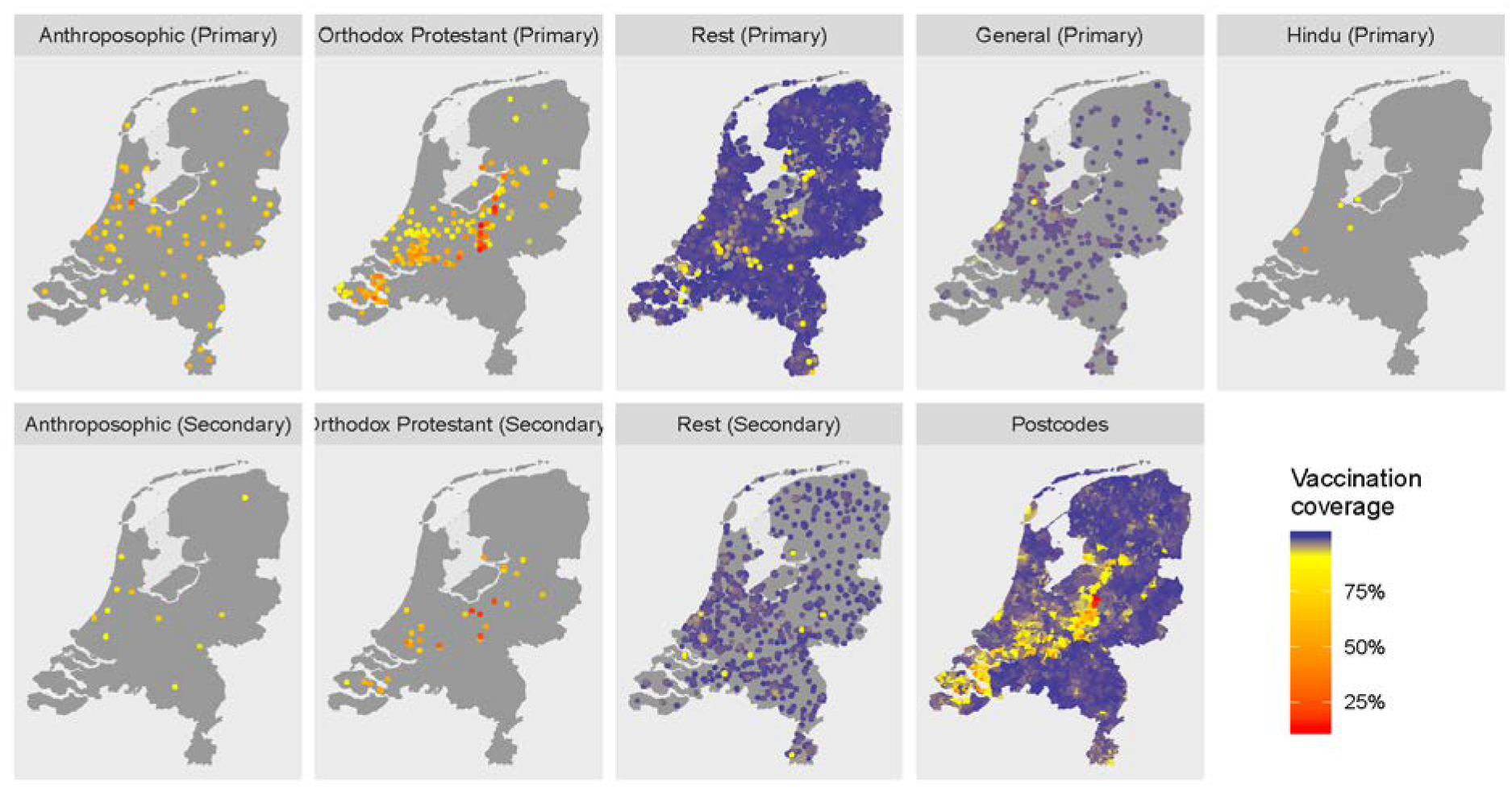
geographic distributions of all schools by identity, with colour coded estimated vaccination coverages. The bottom right map shows the estimated MMR coverages by four-digit postcode area.

Figure 4 shows maps of the school locations, partitioned by school identity, colour coded to indicate estimated vaccination coverage, and as a reference a postcode map of estimated vaccination coverages. The Orthodox Protestant schools are clearly concentrated in the Bible Belt, with most of the very-low coverage schools east from the centre. The lower-coverage Rest schools are also predominantly located in the Bible Belt area; most of these have the Protestant identity (not shown). Anthroposophic schools are more widely distributed across the country.

### Clustering of unvaccinated children

In our age cohort dataset of 197,382 10-year old children in the Netherlands in 2013, 4998 (2.5%) were not vaccinated against MMR. In primary schools, 51% of unvaccinated children attend General or Rest schools most of which have a very high vaccination coverage, but the rest is clustered in schools with much lower coverages: 38% in Orthodox Protestant schools, 10% in Anthroposophic schools, and 1% in Hindu schools (Figure 3, Table 3). When these children go to secondary school, this hardly changes: 56% go to Rest, 39% to Orthodox Protestant, and 5% to Anthroposophic schools. Many children move to secondary schools of the same identity as their primary school, more so in the Orthodox Protestant community (85%) than in the Anthroposophic community (44%), showing that these children go to low-coverage schools during their whole school career. There were, however, no children moving between schools with these two identities.

## Discussion

Our aim was to estimate single-dose MMR vaccination coverages on schools in the Netherlands, and assess clustering of unvaccinated children in schools. It turns out that about half of the unvaccinated children are clustered into a small minority of schools with very low vaccination coverage. These are almost all schools with an Orthodox Protestant or Anthroposophic identity. That implies that thousands of children per cohort are not protected by vaccination, even when the mean vaccination coverage of 97.5% is well above WHO recommendations for both infections.

Spatial clustering of unvaccinated children in the Bible belt is clearly observed at the postcode area level (Figure 4), and has been described before. Our results, however, show that clustering is much stronger at the level of schools, which better reflects the social environment of children. Most schools have vaccination coverages higher than in most postcode areas, whereas some schools of mainly Orthodox Protestant and Anthroposophic identity have much lower coverages (Figure 3). The analysis also revealed social clustering outside the Bible belt area (Figure 4). Apart from the good agreement with the validation data, the Orthodox Protestant coverages also confirm estimates of 60% in this community from online surveys by Ruijs et al [19].

Whereas Orthodox Protestant and Anthroposophical schools are only minority identities, about half of all unvaccinated children attend these schools with low vaccination coverage (Table 3). Children not only spend much of their time at school, their social network outside school is probably very similar. That creates an infection risk for many unvaccinated children, who seem to be protected when considering their residential four-digit postcode area (Figure 5), let alone the national vaccination coverage. However, for virus introductions to cause large epidemics, there should also be sufficient contacts between the schools. Most of these contacts are probably through households, mainly between primary and secondary schools. Aside from the geographic proximity (Figure 2), the school identity careers (Table 2) show that the Orthodox Protestant community is closely knit: 85% of children attending Orthodox Protestant primary schools, also attend secondary school with that identity, and vice versa. In addition, families in the Orthodox Protestant community tend to be large. This explains the occasional large epidemics mostly within this community. Because secondary schools are essential links in the school network, large epidemics may only occur when the interepidemic period is long enough so that these schools are populated with susceptible children. This is consistent with the pattern of measles epidemics in the Bible Belt in the Netherlands, which took place in 1988, 1999, and 2013 [7, 30].

Anthroposophic schools are less well connected, firstly because they are more evenly distributed geographically (Figure 2), and secondly because more children going to either an Anthroposophic primary or an Anthroposophic secondary school but not both (Table 2). Indeed, outbreaks in Anthroposophic schools tend to stay confined to a few schools [31]. Also, the school feeder data suggest that the two low-coverage communities are socially separated, as there were no children in the dataset moving between Orthodox Protestant and Anthroposophic schools (Table 2). As a consequence, during the epidemics in the Orthodox Protestant community, Anthroposophic schools were hardly affected [7].

The estimated school vaccination coverages captured the trend of the validation datasets well, but were too unreliable to report for individual schools (Figure 2). First, they were unprecise, which is reflected by the wide credible intervals, and caused by the large number of schools compared to postcode area coverage data. Second, in a few cases estimates were inaccurate, notably for two schools of the Rest identity (Figure 2), which were Protestant by their registered identity (before aggregation of identities). Misclassification of some Protestant school is likely: the websites of these schools reveal an Orthodox identity, and most low-coverage Rest schools in the Bible Belt (Figure 4) are also Protestant. Third, confounding may have resulted in the estimated low coverages of the Hindu schools, as a lower coverage was not associated with the Hindu religious group in a large cross-sectional survey [32], in which 32 out of 35 Hindu respondents of age 1.5-20 were vaccinated (Mollema, personal communication). Fourth, we used data of a single birth cohort, which may not be representative if the vaccination coverage has increased or decreased, and which may overestimate coverage in younger children if they receive their first MMR vaccination at the age of nine, when the routine second vaccination is offered. If reliable coverages could be obtained, e.g. by coupling of school and vaccination registries, they could be used to inform parents, to prioritize supplemental vaccinations during outbreaks, or for more detailed studies into changes in vaccine hesitancy in different social groups, representated by school identity.

Pockets of low vaccination coverage and resulting risk of outbreaks are better identified in social than geographic environments [33]. We assessed heterogeneity in vaccine coverage by estimating coverages at schools. The reason is that schools are good reflections of the social environment of children, especially if parents can choose a school with a clear identity like in the Netherlands. For the Netherlands, we were able to estimate school-level vaccination coverages by relating available postcode-level coverages to schools through the school catchment data, and we could use school identity to explain much of the variation between schools. In combination with the school feeder data, we could distinguish two well-connected by mutually separated clusters of schools with low vaccination coverage. For other countries, the number of postcode areas may be larger than the number of schools, which would simplify analyses, or alternative variables such as public and private schools may be more useful. Ideally, school vaccination coverage data are directly available: for instance, in the United States these are available at school level for some states [23], but often grouped by county for analysis [26]; also in Germany they are collected at school entry and then grouped by district [24]. For describing connections between schools, feeder data may be supplemented (or replaced) by individual level household data to build larger networks of schools and identify communities at risk.

## Data Availability

Some data are privacy sensitive because of small numbers and may not automatically be shared. Please contact the authors to explore what is possible.

