## Supplemental Methods and Figures for "Social clustering of unvaccinated children: measles-mumps-rubella vaccination coverage in schools in the Netherlands"

These are supplementary methods with the article “Social clustering of unvaccinated children: measles-mumps-rubella vaccination coverage in schools in the Netherlands”

### Supplementary Methods
### Detailed methods and preliminary analyses

#### Datasets and model input

We used five datasets to estimate school vaccination coverages (dark green in Figure 1, main text). These datasets were used to build the population structure, and the actual model input (light green in Figure 1, main text). We used two datasets for validation of estimated coverages (Utrecht and Anthroposophic school data).

##### Datasets for estimation

Vaccination coverage data **Y, N** came from the Dutch vaccination registry Praeventis [1]. The MMR vaccine is given at 14 months and at 9 years of age, and coverage data were available for 2-year olds and 10-year olds. We used the coverage in 2013 of at least one vaccination at 10 years of age (birth cohort 2003), which will best describe how well protected the schools are. Cohort coverage data per four-digit postcode $i$ are the number of vaccinated children $y_i$ and the total number of children $n_i$ in that age class. The dataset contained 197,382 children in 3845 postcode areas.

School attendance data came from the Education Executive Agency of the Ministry of Education (DUO) [2]. We used the following datasets:

- the primary school catchment data **P**: residential postcodes of primary school children of eight years and older in 2012, with $P_{ij}$ being the number of children in postcode $i$ attending school $j$. This dataset contained information about 830771 children at 7570 schools.
- the secondary school catchment data **S**: residential postcode of secondary school children in the first three grades in 2013, with $S_{ik}$ being the number of children in postcode $i$ attending school $k$. This dataset contained information about 648606 children at 1770 schools.
- the school feeder data **F**: children moving from primary to secondary school in September 2013, with $F_{jk}$ being the number of children going from primary school $j$ to secondary school $k$. This dataset contained information about 205606 children.
- school identity data **D**: schools were categorised into 15 groups based on their identity. For each primary school $j$ and secondary school $k$, identities $D_j^P$ and $D_k^S$ were registered. A special type of identity was formed by all collaborative identities (“Samenwerking”), which consisted of schools in which two or more identities collaborate in several combinations of Protestant, Roman Catholic, General, and/or Municipal. All unspecified identities in secondary schools (categorized as Rest (“Overige”) in the dataset, or left blank) were treated as a separate identity “Rest”.

The school database changes every year, so we chose to use two different years for the primary and secondary school residential data to maximize the consistency with the school feeder data. Schools that were not present in the school feeder data (26/7570 primary and 5/1770 secondary schools) were added to these data, each linked to a unique dummy school of the same identity. Reasons for missing were that schools were new or very small.

The reason not to include children younger than eight years of age, is that before eight years there is still a considerable flow from regular primary schools to special schools for children needing extra attention (“Speciaal Onderwijs”). The reason not to include children above third grade is that many children leave secondary school after that grade. Between eight years of age and third grade, most children attend one primary and one secondary school, which will be referred to as their ‘school career’.

##### Datasets for validation

Validation data came from two sources

- **Utrecht schools dataset**: collected by the municipal health service Utrecht, covering part of the Bible Belt. They collected school-level vaccination coverages during the 2013 measles epidemic by asking schools for student lists and coupling these to the vaccination registry Praeventis. This resulted in coverages for 488 schools.
- **anthroposophic school dataset**: collected during a study in 2014 on vaccination hesitancy on anthroposophic schools, resulting in publication of coverage level for 9 primary and 2 secondary schools [3].

**Model input for estimation**

In the analysis, we modelled the vaccination status of children (observed per postcode) as a function of their school career, so we needed the numbers of children in each postcode *i* with school career $\{j, k\}$:

- school career population structure **C**: numbers of children $C_{ijk}$, living in postcode *i*, attending primary school *j* and secondary school *k* during their school career. These were calculated with datasets **P**, **S**, and **F**:
  1. the number of children $P_{ij}$ that live in postcode $i$ and go to primary school $j$, are distributed across secondary schools $k$ proportional to the product $S_{ik} \cdot F_{jk}$:
  $$P_{ijk} = P_{ij} S_{ik} F_{jk} \Big/ \sum_{k'} S_{ik'} F_{jk'} \tag{1}$$
  2. the number of children $S_{ik}$ that live in postcode $i$ and go to secondary school $k$, are distributed across primary schools $k$ proportional to the product $P_{ij} \cdot F_{jk}$:
  $$S_{ijk} = S_{ik} P_{ij} F_{jk} \Big/ \sum_{j'} P_{ij'} F_{j'k} \tag{2}$$
  3. finally, the school career population structure $C_{ijk}$ is calculated as the sum of $P_{ijk}$ and $S_{ijk}$:
  $$C_{ijk} = P_{ijk} + S_{ijk} \tag{3}$$

For the analysis, the vaccination coverage for each school career is described by three parts:

- part 1 explained by the identity career **X**, which indicates the two identities of the schools *j* and *k*
- part 2 explained by the primary school $\mathbf{Z_1}$, which indicates the primary school *j*
- part 3 explained by the secondary school $\mathbf{Z_2}$, which indicates the secondary school *k*

#### Statistical analysis

Our aim was to estimate MMR vaccination coverage by school. We made the assumption that the coverage data for the 10-year olds is representative for coverage in other age cohorts. That made it possible to express the vaccination coverage per postcode area in terms of the school careers of the children in that postcode area.

*General regression model*

In our model, we described the vaccination coverages per postcode in terms of the school careers attended in that postcode, and fitted these coverages to the postcode-level coverage data:

$$y_i \sim \text{Binom}(n_i, p_i) \tag{4}$$

Here, postcode coverage $p_i$ is a function of the data and model parameters, together describing the contributions of the vaccine coverages per school career $v_{jk}$ to the postcode vaccination coverages:

$$p_i = \sum_{j,k} C_{ijk} v_{jk} \Big/ \sum_{j,k} C_{ijk} \tag{5}$$

. The dataset contained more schools than postcode-level coverage data. Therefore, we used hierarchical models with the identities of the two schools in the school career (identity career) as a grouping variable.

Thus, the logit-coverages per school career were modelled by a sum of three parts: a mean depending on the identity career **X**, a deviation for the primary school $\mathbf{Z_1}$, and a deviation for the secondary school $\mathbf{Z_2}$:

$$\text{logit}\left(v_{jk}\right) = \mu_{jk} + d_j^P + d_k^S \text{, with} \\ d_j^P \sim N\left(0, \sigma_{D_j^P}\right) \text{ and } d_k^S \sim N\left(0, \sigma_{D_k^S}\right) \tag{6}$$

We used $\mu \sim N(0,10)$ and $\sigma \sim Cauchy(0,2)$ as prior distributions for all $\mu$'s and $\sigma$'s.

*Preliminary analyses and final analysis*

Because some identities had only few schools, and the total number of unique identity careers would be very large, we wanted to group all identity careers for which there was no evidence of having a lower coverage than most schools. This grouping was done in two steps, in preliminary analyses:

1. Two separate models were run for all primary or all secondary schools, with all identities, so $p_i$ in Eq (5) is expressed in terms of primary school attendance or secondary school attendance only, not the school career. From the sampled posterior, we calculated for each identity the posterior probability P that its mean coverage is lower than the mean coverage of any of the major identities Municipal, Protestant, or Roman Catholic. All identities without evidence of lower coverage (P < 0.90) were grouped into one Rest identity. Results are listed in Table S1. This resulted in five primary school identities and three secondary school identities.
2. A single model for all primary and secondary schools, with all 5 x 3 = 15 possible identity careers from the reduced sets of identities from the first preliminary analysis. In this model, $p_i$ in Eq (4) is expressed in terms of school career attendance per postcode area, as described above (Eq. (5) and (6)). From the results, we calculated for each identity career the posterior probability P that its mean coverage is lower than the mean coverage of the Rest-Rest career. All identity careers without evidence of lower coverage (P < 0.90) were grouped with socially most similar identity careers (one of the two identities changing to Rest). Results are listed in Table S2. This resulted in eight remaining identity careers.

The final analysis (Results in main text) was done with the model in Eqs (4), (5), and (6) as well, but with only the eight remaining identity careers. All models were fitted in a Bayesian framework in stan (mc-stan.org), called from R statistical software with the Rstan package (mc-stan.org/rstan). We ran 10 chains with 2000 samples, of which we removed the first 1000 as warm-up.

**Supplementary Table 1. Numbers of primary and secondary schools of all identities, and posterior probabilities of lower vaccination coverage than the major identities (P > 0.9 in bold)**

| Identity | Primary schools | | Secondary schools | |
|---|---|---|---|---|
| | *n* | Pr(lower coverage) | *n* | Pr(lower coverage) |
| Municipal | 2444 | reference | 352 | reference |
| General | 497 | **0.92** | 392 | 0.11 |
| Anthroposophic | 70 | **1.00** | 11 | **1.00** |
| Roman Catholic | 2236 | reference | 248 | reference |
| Protestant | 1882 | reference | 281 | reference |
| Reformed (Liberated) | 118 | 0.02 | 9 | 0.08 |
| Orthodox Protestant | 180 | **1.00** | 34 | **1.00** |
| Evangelic | 13 | 0.19 | 3 | 0.20 |
| Evangelical Fraternity | 2 | 0.36 | 0 | - |
| Hindu | 6 | **0.93** | 0 | - |
| Islamic | 43 | 0.51 | 1 | 0.47 |
| Jewish | 2 | 0.61 | 2 | 0.58 |
| Interconfessional | 12 | 0.05 | 4 | 0.12 |
| Collaborations | 65 | 0.58 | 362 | 0.32 |
| Rest | 0 | - | 71 | 0.24 |

**Supplementary Table 2. Numbers of children in primary-to-secondary school feeder data by school identity career, and posterior probabilities of lower vaccination coverage than the Rest-Rest career (P > 0.9 in bold)**

| Primary school identity | Secondary school identity | | | | | |
|---|---|---|---|---|---|---|
| | Anthroposophic | | Orthodox Protestant | | Rest | |
| | *n* | Pr(lower coverage) | *n* | Pr(lower coverage) | *n* | Pr(lower coverage) |
| Anthroposophic | 679 | **1.00** | 0 | - | 876 | **1.00** |
| Orthodox Protestant | 0 | - | 4298 | **1.00** | 765 | **1.00** |
| Hindu | 0 | - | 0 | - | 262 | **0.91** |
| General | 182 | 0.41 | 3 | 0.75 | 13246 | **0.96** |
| Rest | 778 | 0.30 | 782 | **1.00** | 182471 | reference |
